# Clinical Decision Support Systems implementation in Africa: a systemic review

**DOI:** 10.1101/2023.10.07.23296693

**Authors:** Jacob Nii Noye Nortey, Kate Takyi, Andrew Adabo, Rashida Suleiman

**Affiliations:** Department of Computer Science, Kwame Nkrumah University of Science and technology

**Keywords:** Clinical decision support systems, Africa, health informatics, and decision support systems

## Abstract

The healthcare system in Africa is characterized by the lack of trained clinicians, resources, brain drain and quality care processes. To mitigate these challenges, some African countries have resorted to the use of health information technologies such as the Clinical Decision Support System (CDSS). Globally, CDSS implementations in the health sector have been reported to have reduced medical error, increased hospital accessibility and standard of care. Studies on the number of implemented CDSS in Africa were found to be limited since only a few are available. Despite this setback, the few implemented CDSSs are effective, efficient, and reliable in the diagnosis of diseases. In order to fully ascertain the impact of CDSS implementation in African counties, we evaluated the documented CDSS programs that are in operation. An extensive search was performed on Google Scholar, PubMed, and SCOPUS. About 38 (thirty-eight) publications were identified, of which some met the inclusion criteria. Limited implementation of CDSS was observed in the African countries. The review showed promising impacts of CDSS in African healthcare centers especially in the diagnose and treatment of pediatric and maternal related disease.

## 1.0 Introduction and Background

It is approximated that more than eight million people from Low - and Medium - Income Countries (LMIC) die annually due to diagnosis error, poor healthcare quality and medical error [1]. Consequently, in 2015, an estimated amount of $6 million was lost due to decreased productivity [1]. Studies conducted in the United States indicated that up to 9800 US citizens die annually due to avoidable medical or diagnosis error [2]. Schiff et al. defined diagnosis error as inaccuracy or aberrations in medical diagnosis procedures leading to over diagnosis, under-diagnosis, or concealed diagnosis [3]. Another study conducted in the US on the topic “errors of diagnosis in pediatric practices” attested that 54% of the pediatricians indicated making diagnosis errors at least twice a month [4].

To address these setbacks, countries around the world, especially the developed countries, have made huge investments in the health sector [5]. One of such investments is in the usage of health information technology to achieve quality healthcare. This paper seeks to evaluate the implementation of one of the health information technologies; Clinical Decision Support System (CDSS) in Africa. Traditionally, CDSS was designed as part of the Computerized Physician Order Entry (CPOE) application and functions as a device used to prevent over diagnosis. However, Kawamoto et al, defined CDSS as a program module designed to help clinicians make decisions about a patient at a point of care [5]. CDSS over the years is useful in the diagnosis, management, and treatment of diseases.

Many health information management organizations have developed and implemented numerous clinical decision support systems over the past decades. A typical example of a CDSS is DXplain. This decision support system was originally developed by Dr. Barnett Octo as an education tool [6]. It is now used commercially for both educational and diagnosis purposes. Another example is the Theradoc antibiotic assistant CDSS. This tool combines a patient’s electronic medical history, laboratory investigations, pharmacy, and radiological sections to prescribe an antibiotic administration to a patient [7]. Similarly, MYCIN and Firstline decision support systems also work in this manner to provide effective antibiotic choice to patients. Other examples of CDSS include; EuroCAT, HELP, and Quick Medical Reference, etc.

## 2.0 Literature review

The healthcare system in Africa is characterized by poor quality of service, lack of equipment, and deficits in the number of health practitioners. Oleribe et al. research in Mozambique identified challenges with human resources, financial quotas to health services, and poor management services [8]. Similar studies by Edzie in Ghana and Nigeria also observed a huge deficit in the number of radiologists per 100,000 patients, which is more than WHO recommendations [9]. In effect, one can further conclude that the healthcare system in Africa, especially in Sub-Saharan African countries, is a weak system, which is a major barrier to fulfilling the Sustainable Development Goals such as SDGs 3, 4, 5, and 6 [10].

### 2.1 Clinical Decision Support Systems in Africa

Literature reviews indicate a lack of CDSS implementation in Africa. This may be because of the lack of funding in the health sector [8]. Another reason may be the lack of robust health information management system technologies on the continent. Health information technologies are useful in the design and implementation of any CDSS. This is because it provides an overview of patient information, which forms the backbone in the design of CDSS [11].

Furthermore, some African countries have still not implemented the electronic medical record system and are using the conventional methods. This smacks down on the design and implementation of CDSS. Despite all these challenges, some African countries are making progress in the gradual rollout of decision support systems. There have been many pilot programs in most of these African countries to evaluate the efficiency of CDSS in mitigating some of the health service challenges. Most of these pilot programs are largely driven by HIV treatments and maternal and neonatal mortality [12,13].

### 2.2 Functional modalities of Clinical Decision Support Systems

#### 2.2.1 Cost containment

Cost containment can be defined as the process of effectively cost reductions in business. When implemented strategically, it can help increase the profit of an organization. Cost containment in CDSS is very important to both the patient and the health care provider. Research conducted by Algaze et al. found a reduction in the usage of laboratory resources when Computer Order Entry (CPOE) and daily checklist rule were used without affecting mortality rate [14].

Laboratory reagents are costly to the health care provider and the laboratory test is also costly to the patient. Therefore, with the advent of CDSS, we can eliminate tests, which are irrelevant to the diagnosis since the CDSS has a high prediction rate. Again, with this system, we can save money and time to both the patient and hospital.

Additionally, another research by McMullin et al. also showed the reduction in the cost of prescription by $4.99 in the interference group [15]. CDSS can also provide cheaper alternatives to drugs. In Germany for instant, research showed 93.6% of drugs switch to cheap alternative drugs [16]. One of such CDSS system tools is MYCIN, which gives therapeutic advice and provides a cheap alternative to each case.

#### 2.2.2 Administrative functions

CDSS tends to increase hospital documentations. Documentation of electronic health records is important for the early diagnosis of certain disease types. Documentation of splenectomy was also observed to have increased when the CDSS was implemented [17]. This documentation was performed to monitor patients who have been vaccinated following splenectomy, which has increased the risk of infections. Again, the implementation of CDSS was also observed to have addressed the inconsistencies and inaccuracy of coding of the International Statistical Classification of Disease (ICD) [18].

#### 2.2.3 Diagnostic support

Clinical decision support systems are computed schemes which are designed to aid clinicians in diagnosis. CDSS mainly designed for diagnosis purposes are called Diagnosis Decision Support (DSS) and an example is Dxplain. Research conducted by Porat et al. reported that 74% of general practitioners found the Diagnosis Decision Support useful [19]. The general practitioners also said that the DSS enabled them to broaden their extent of diagnosis. Another study by Kunhimangalam et al. also observed 93% accuracy in the diagnosis of peripheral neuropathy [20].

#### 2.2.4 Managing clinical complexity and details

In most hospitals, disease treatment guidelines are not strictly adhered to. Most general practitioners rely on experience rather than following protocols and guidelines established by health organizations in disease treatments and management. With the advent of CDSS, the protocols have now been adhered to since the guidelines are infused in the design process of the CDSS tools. An example of this is an observational study by Kwokk et al. The study took 50 patients who were then compared with 50 historical bridles. A CDSS tool Asthma Clinical Assessment Form and Electronic decision support (ACAFE) was used to document and manage asthma patients. Statistically significant (p < .01) association with higher levels of certification of severity of asthma was observed because the ACAFE contained guidelines for asthma management which were strictly adhered to compared to the 50 historical controls [21].

### 2.3 Challenges of Clinical Decision Support Systems

Despite all promising benefits of CDSS, the challenges associated with its usage cannot be overridden. A challenge identified by Antoniadi et al is the issue of trust with the system. Clinicians find it difficult to use the CDSS as in certain conflicts with the diagnosis of the attending physician [22]. Another challenge is the perception of general practitioners. Porat et al. reported that 38% of general practitioners felt that the system took a longer time to give an output [19].

Sittig et al. also argued that for an effective implementation and usage of CDSS, the following must be taken into consideration: improve the usefulness of clinical decision support interventions (which includes human – computer interface, prioritization of filter recommendation, usage of free text information etc.), creation of new clinical decision support interventions, and sharing of data driven from CDSS implementations [23].

### 2.4 Ethical concerns with Clinical Decision Support Systems

The usage of technology in healthcare delivery is on the ascendancy, and as such prudent measures must be taken to ensure the safety of patients since CDSS in most countries has been integrated with patients’ electronic health records. Research conducted by Čartolovni et al. concurred that a review of 1,108 papers showed prevalent issues with patient protection, liability and accountability on both the CDSS developer and the healthcare center, lack of meaningful documentation and regulations and the effect of CDSS on the patient and physician relationships [24]. One can therefore argue that in any event of wrong medical diagnosis of the decision support system, which leads to the death of the patient, who becomes liable and who will be accountable.

Another research conducted by De Panfilis et al. also illustrated the need for the right to autonomy by patients when an artificial intelligence-based decision support system is used to predict palliative care. CDSS tends to predict and signal treatment to patients who require extensive and complex treatments [24,25]. It is necessary to take into consideration the emotional state of the patient, financial, and right to quality of life. Therefore, after the decision support system has indicated the treatment plans, steps must be adopted to take into consideration the rights of the patient.

## 3.0 Method

### 3.1 Publication identifications

Thorough searches were made on Google Scholar, SCOPUS, and PubMed employing the following key terms;

- Clinical decision support system and Africa
- Clinical decision support systems and implementation
- Health informatics and Africa
- Decision support system and Africa

### 3.2 Inclusion criteria

Publications of interest, which were in line with this study’s literature, were selected. A total of eight (8) publications were obtained.

### 3.3 Exclusion criteria

Reviews, duplications, and articles that do not fit literature review and personal opinions were eliminated. Fig 1.0 indicating evidence of search

**Table 1.**
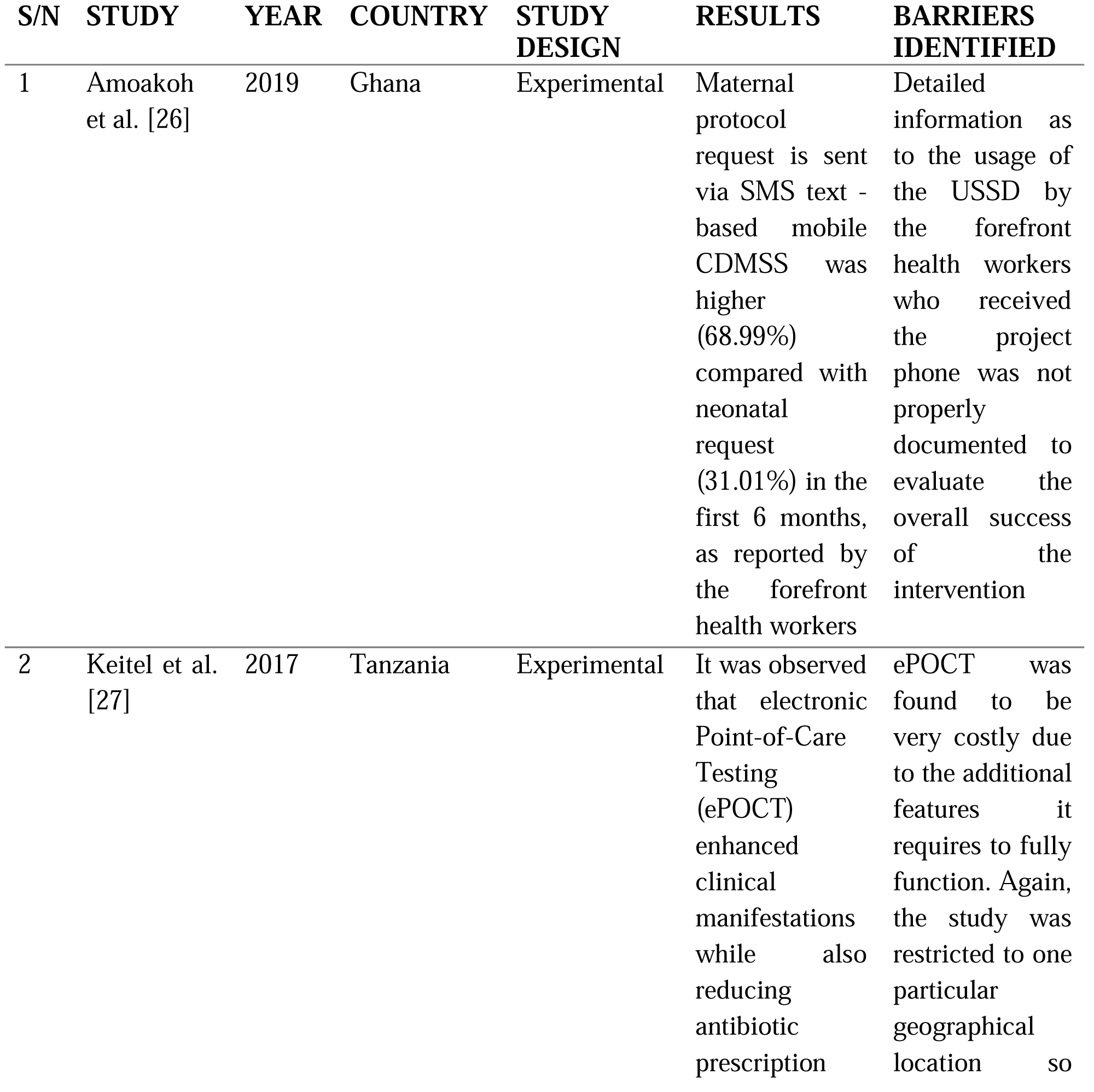
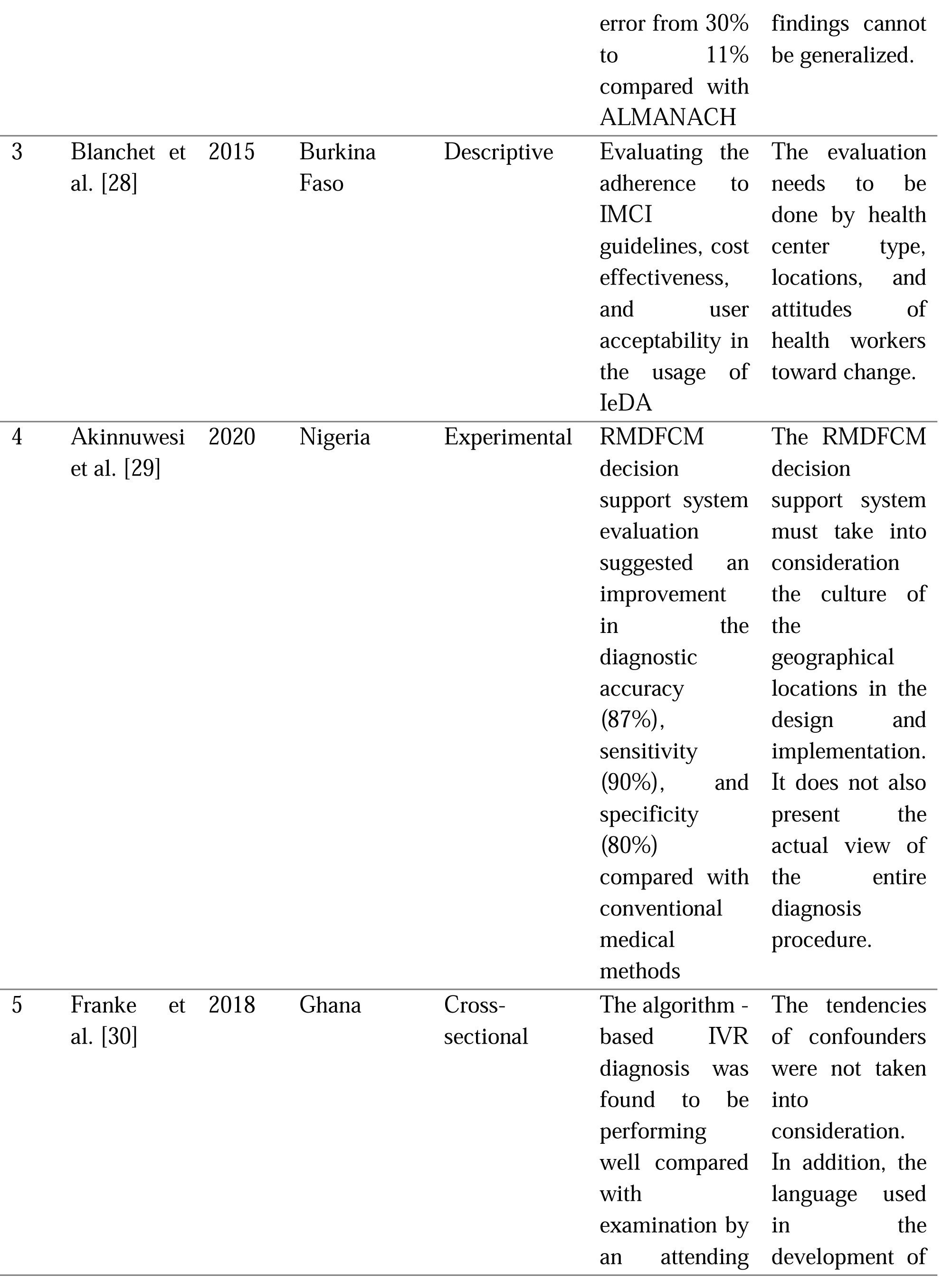
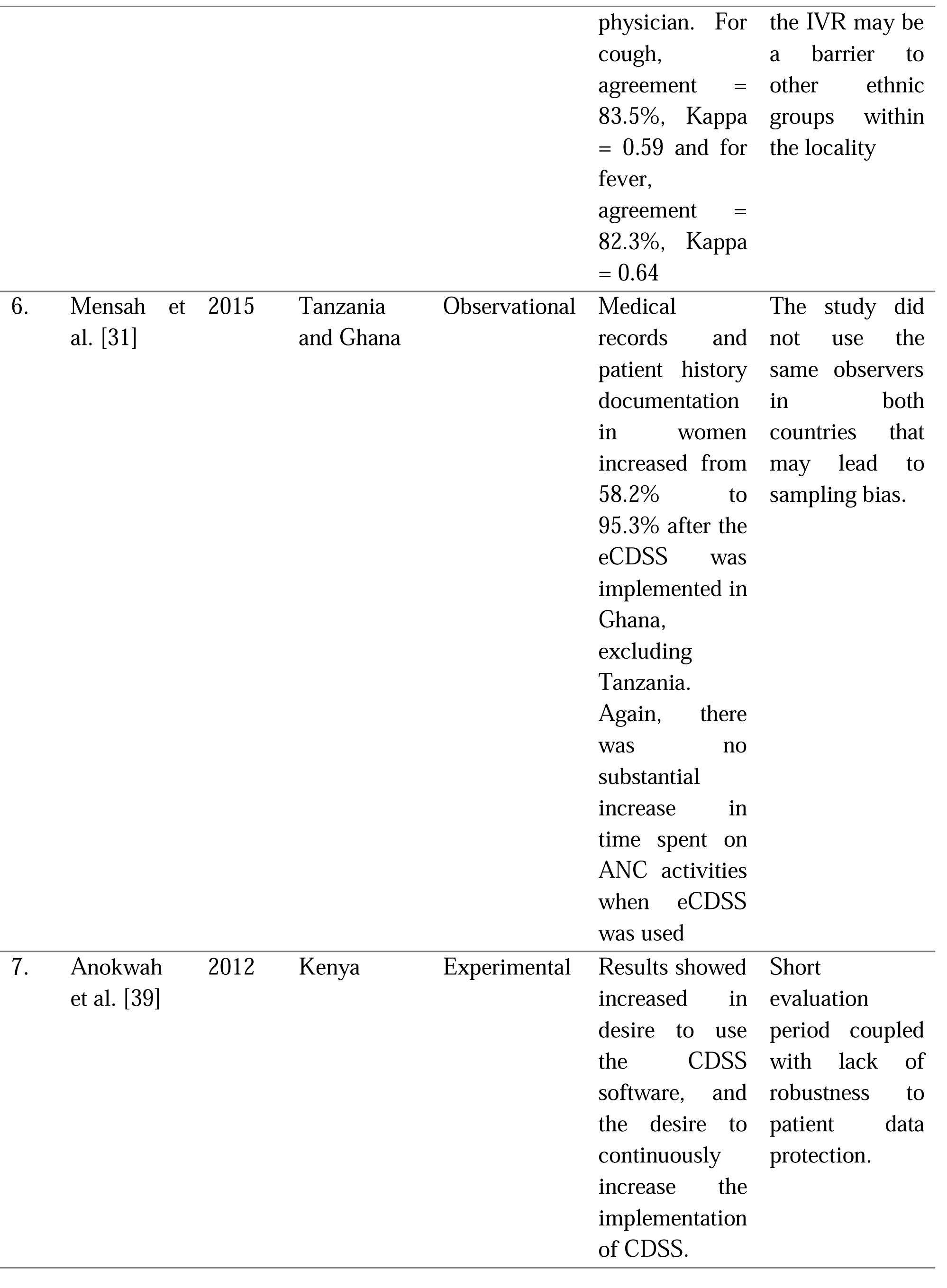
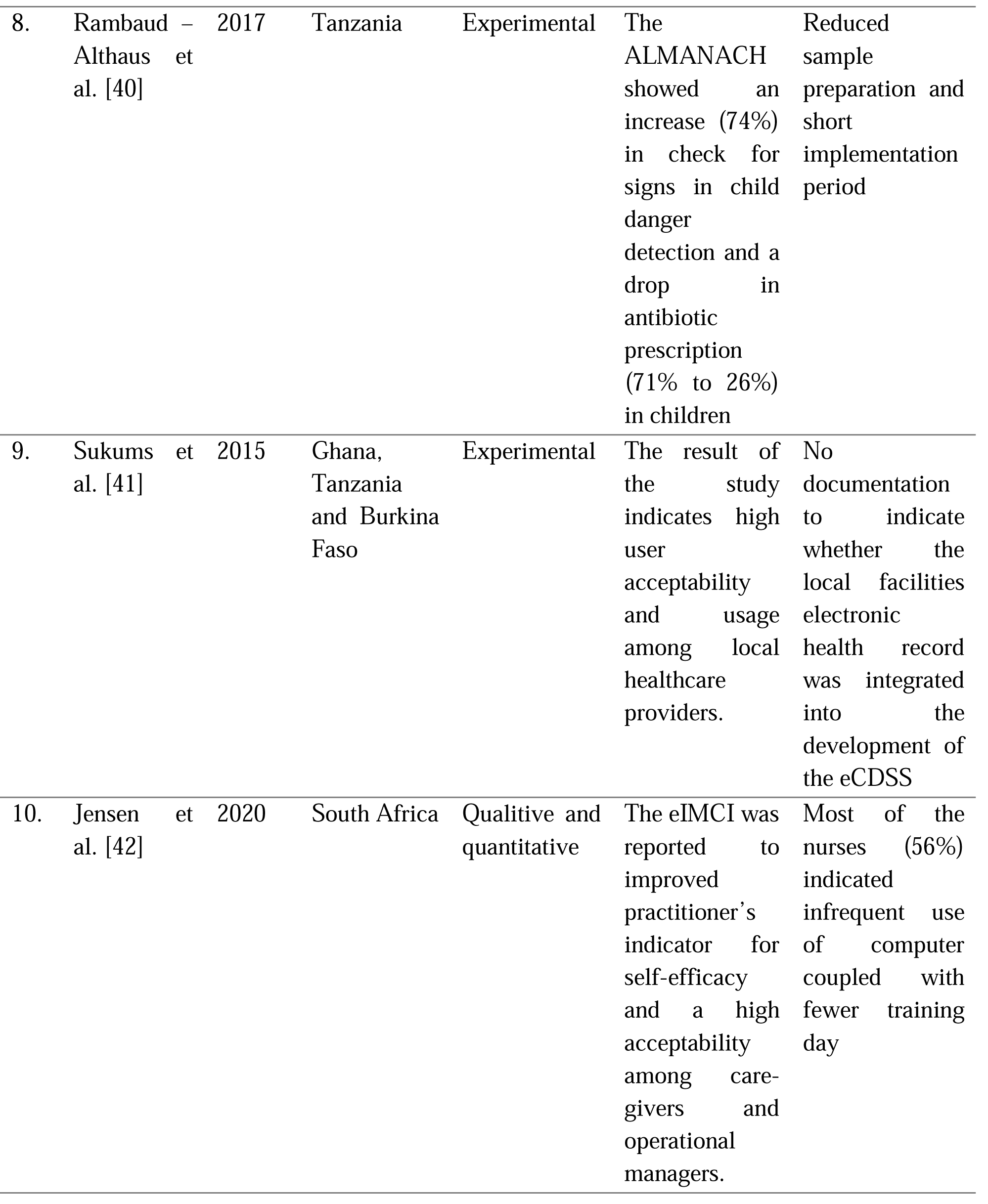
A summary of research attempting to quantify the scope and success of implemented clinical decision support systems in Africa.

## 4.0 Discussions and Findings

Literature search on the implementation of clinical decision support systems in Africa is limited [26-31]. This may be due to financial constraint, lack of proper documentation of implemented decision support systems, lack of training in health informatics, and the failure of countries to share data driven from the implementation of clinical decision support systems in Africa [32-34]. In effect, the analysis of the statistical usage and impact of the implemented CDSS cannot be fully ascertained to make meaningful policies. Discussions on the implemented CDSS in this paper have been categorized into three: (a) reliability, accuracy and speed of the implemented CDSS, (b) cost effect of CDSS, and (c) cultural integration of CDSS in healthcare centers in Africa.

### 4.1 Reliability, accuracy, and speed of implemented clinical decision support systems

To successfully implement CDSS, speed, accuracy, and reliability are very crucial. Regardless of how wonderful a decision support system is, if it is slowing the workflow or takes lot of time to produce a report, it will be worthless and ineffective [35]. Lee et al. study on “implementation of physician order entry: user satisfaction and self – reported usage pattern” indicated that the ultimate determinant of user satisfaction in electronic CDSS is speed [36]. It was observed in the review of the Mensah et al. study that there was no significant increase in the time spent at ANC when the CDSS was implemented. Workflow was also found to be running smoothly without any delay [31]. Similar studies by Overhage et al. also argued that most of the physicians attested to the fact that the implemented CDSS improved their patient care [37].

Additionally, Akinnuwesi et al. observed the accuracy and reliability of their implemented decision support system. The implemented RMD FCM decision support system improved the diagnosis accuracy (87%) and sensitivity (90%) [29]. Again, a review of Franke et al. study in Ghana also indicated that the diagnosis made by the algorithm - based IVR was similar to attending physician examinations (for cough: agreement = 83.5%, Kappa = 0.59, for fever: agreement = 82.3%, Kappa = 0.64) [30]. In view of this, one can therefore argue that the CDSS program implementation in Africa has shown on various levels that it can be accurate and reliable in the diagnosis of disease.

### 4.2 Cost effectiveness of a clinical decision support system

In any project management, cost effectiveness needs to be assessed to obtain the value of the project against the cost. The study by Keital et al., on the implemented ePOCT was found to be costly since it incorporated additional relevant features to the POCT [27]. It was observed from literature reviews that, there is limited data on the cost analysis of CDSS implementations in Africa. Future studies must focus on the cost analysis of CDSS.

### 4.3 Cultural integration of Clinical Decision Support System

The success of decision support systems can also be attributed to the proper integration of the healthcare center’s cultural practices in the design processes. Cultural integration is defined as the ability of CDSS algorithms, alerts, and guidelines to fit into the workflow of clinicians [38]. One limitation in the study by Blanchet et al. was the inability of the implemented decision support system to take into consideration the attitudes of healthcare workers toward change and the local needs of the individual healthcare centers [28]. It is therefore important to solicit the views, policies, and workflow of healthcare centers and integrate them into the algorithm of the CDSS. It is also important to note that the development of CDSS must not be generalized, and it should be planned to meet the specific needs of individual healthcare centers to achieve the desired results.

## 5.0 Conclusion and recommendations

The implementation of Clinical Decision Support Systems in various African countries is promising. It has been demonstrated that it can be reliable and accurate in the diagnosis of diseases. This will help mitigate the compounding problems of health sector brain drain and lack of quality healthcare practices. The limitation of this study is the ability to provide more evidence on the cost analysis of the implemented CDSS in African countries.

In order to actualized the potentials of clinical decision support systems in Africa, we recommend the following; (1) Future studies should focus on comprehensive evaluation of cost analysis of CDSS implementation in Africa. (2) Health education institutes must include the concept and practical teaching of CDSS in their curriculum. (3) African countries must establish, enforce and regulate laws concerning accountability and liability in the development and implementation of CDSS.

## Data Availability

All data produced in the present work are contained in the manuscript

## Notes

### Competing Interest Statement

The authors have declared no competing interest.

### Funding Statement

This study did not receive any funding

