## Supplementary figures and images for "Clinical Decision Support Systems implementation in Africa: a systemic review"

### supplementary fig 1.0

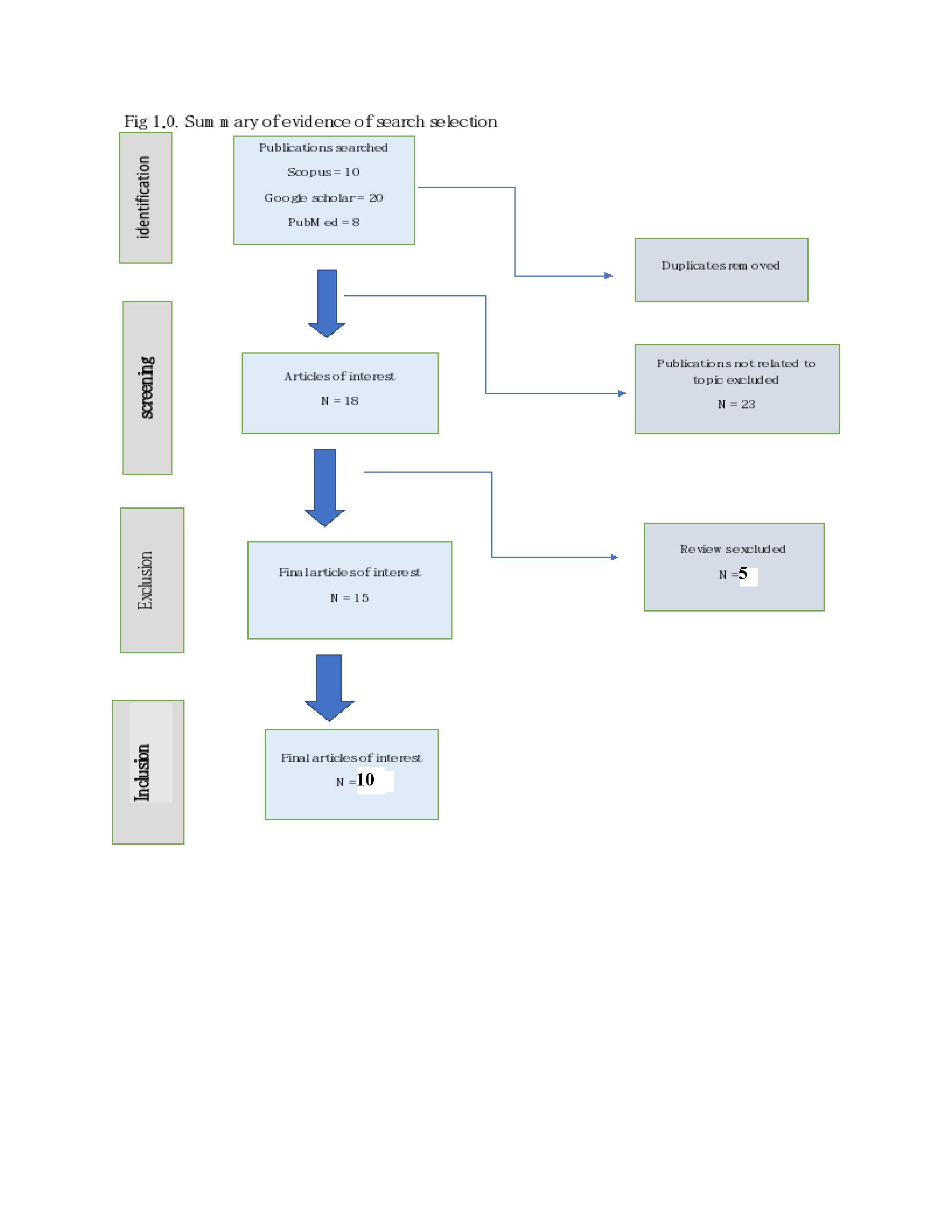
